# Understanding potential non-malarial benefits of sulfadoxine-pyrimethamine treatment during pregnancy on birthweight: a scoping review

**DOI:** 10.1101/2025.05.24.25328278

**Authors:** Swapnoleena Sen, Pablo Martinez de Salazar, Joerg J. Moehrle, Melissa A Penny

## Abstract

**Background:** Malaria in pregnancy affects both maternal and infant health. The World Health Organization recommends administering at least three doses of intermittent preventive treatment (IPTp) using sulfadoxine-pyrimethamine (SP) in *P. falciparum* malaria endemic areas during the 2^nd^ and 3^rd^ trimester of pregnancy. Recent clinical trials have described antimalarials with superior antimalarial effects in the mother compared to SP but led to inferior impacts on infant health outcomes such as low birthweight. Secondary non-malarial effects of SP are postulated to contribute to foetal growth and infant health; however, these remain poorly defined. In this scoping review, we aimed to improve the current understanding of the overall prophylactic effects of antimalarial drug use in pregnant women.

**Methods:** A systematic search using PubMed, Embase and CENTRAL databases in May 2024 was conducted in accordance with PRISMA-ScR guidelines. Results from randomized controlled trials, as well as observational studies, pre-clinical studies, and meta-analysis published between 2004 to 2024 were extracted. Search terms included “malaria” and “pregnancy” and “inflammation” or “angiogenesis” and “birth” in titles or abstracts. The search strategy was expanded to exclude “malaria”, and to include “birthweight” or “birth outcome”. Studies were included if inflammatory and/or placental angiogenesis biomarkers and birthweights were reported.

**Results:** Following a blind review of 111 articles by two reviewers, 12 were included to chart key results. Three additional studies were included by citation and website search. The results indicated that the potential non-malarial benefit of SP may primarily include: i) reduction of systemic or off-target local inflammation or ii) positive regulation of placental angiogenesis or both. Data gaps were identified and addressed as several action measures proposed for consideration in future studies.

**Conclusion:** Characterization of potential off-target effects of IPTp-SP on improving birthweight could include biomarker data for inflammation, and placental angiogenesis, categorized by gravidity and at multiple time points throughout the chemoprevention period. Inclusion of such data in future empirical studies is anticipated to facilitate our understanding and estimation of the overall public health impact of current IPTp drugs. This could aid clarification of concepts of secondary benefits to support informing preferred product characteristics for IPTp candidates.

## Background

Malaria in pregnancy (MiP) continues to be a major public health burden affecting more than one-third of pregnant women in malaria endemic regions of Africa[1]. Each year, it is reported that approximately 75,000–200,000 preventable infant mortalities are attributable to severe foetal and neonatal outcomes as a result of MiP caused by *Plasmodium falciparum* (*P. falciparum*) parasites[2] The risk is particularly high for first and second pregnancies (primigravidity and secundigravidity) due to insufficient protective antibody build up and reduces gradually in those with subsequent pregnancies (multigravidity)[3, 4]. To address this, the WHO recommends prophylactic intermittent preventive treatment in pregnancy (IPTp) with at least three doses of effective antimalarial drugs to first- and second-time mothers (primigravidae and secundigravidae) living in *P. falciparum* endemic settings[5]. Currently, sulfadoxine-pyrimethamine (SP) is given during their antenatal care (ANC) visits at their second and third trimester of pregnancy[5].

Treatment with SP remains effective in settings with partially resistant parasites, although with gradually shortened prophylactic periods depending on type of mutation[6, 7]. To combat the potential loss of adequate protection with increasing parasite mutations, efforts have been made to explore alternative IPTp drug candidates[8, 9]. This has led to the identification of novel candidates and interventions with desirable properties, such as, longer duration of protection and single dosing schedule[10–12]. Notably, these explorations have raised the importance of also evaluating non-inferiority traits for all treatment options, as well as a re-evaluation of the impact and mode-of-action of SP[7, 13, 14].

Findings from several clinical trials[15–19] and a mediation analysis[20] have shed light on this issue. For example, one clinical trial assessed the impact of IPTp with dihydroartemisinin-piperaquine (IPTp-DP) with additional antibiotic azithromycin (AZ) compared to IPTp-SP[18, 19]. Neither IPTp-DP nor IPTp-DP+AZ could improve infant health outcome, particularly low birthweight (LBW) compared to IPTp-SP. In addition, a separate trial found reported symptomatic malaria and peripheral parasitaemia following treatment with IPTp-AZ plus chloroquine (IPTp-AZCQ) to SP. However, IPTp-AZCQ showed non-superior birthweight measures[17]. Furthermore, since IPTp-SP is contraindicated for patients receiving anti-retroviral therapy, one trial assessed an alternative antimicrobial, Mefloquine (MQ), for IPTp use case and found reduced maternal parasitaemia at delivery in IPTp-MQ recipients but non-significant benefit against LBW[21].

It is hypothesized that the total impact of IPTp-SP extends beyond its anti-malarial benefit, and such putative non-malarial effects may also contribute to foetal and infant health outcomes[15–19, 22]. There are several assumptions regarding the mechanism of action for such “secondary effects” beyond the broad spectrum antibiotic effects of sulfadoxine[14, 18, 19], as well as the anti-inflammatory effects of pyrimethamine[23, 24]. Surprisingly, findings from the trial comparing IPTp-AZCQ vs. SP[17] reported a higher percentage of maternal co-infection burden in the SP cohort (except for *Chlamydia trachomatis),* but the risk of LBW was lower. Such a finding suggested that the benefits of SP on birthweight could involve other pregnancy-specific pathways. Furthermore, the benefit against LBW may be mediated at least partly by improving maternal microbiota and gestational weight such that lowering the burden of infection and inflammation increases the availability of energy and nutrients for both mother and foetus through shifts towards beneficial bacterial species without dietary changes[18].

A systematic review by Olaleye and colleagues[25] examined randomized or quasi-randomized (i.e. where the participants are allocated to study arms without randomization) controlled trials investigating IPTp with SP vs. different regimens of DP (three-dose or monthly or as intermittent screening and treatment) to examine different IPTp drug effects. This study focused on meta-analysis of the differential impact on maternal malarial parasitaemia and infant health measures. However, potential underlying pathophysiological pathways were not explored.

Collectively, these lines of evidence indicate that there are opportunities to better understand and define the non-malarial effects of IPTp-SP, and to explore how LBW risk and chemoprevention effects are documented as a function of gravidity. Here, we have conducted a scoping review to refine our understanding of the effects of IPTp-SP and propose a roadmap to define its therapeutic properties for mothers and infants as a comparator for alternative IPTp candidates. Our findings are relevant to the evaluation of optimal chemoprevention choices that protect these vulnerable populations against malaria and associated health risks.

## Method

### Study design

We undertook a systematic scoping review to examine a diverse type of study designs (randomized controlled trials, longitudinal observations, and a systematic review)[26]. We chose this approach of evidence synthesis to: i) clarify the concept of non-malarial benefits of antimalarial drugs used in IPTp, ii) narrow down our understanding of the potential pathways through which IPTp-SP may exert such off-target effects, and iii) compile current knowledge gaps that can inform potential action measures for future research [27]. This study was performed in accordance with PRISMA-ScR guidelines[28].

### Information sources

This study focused on extracting empirical evidence from peer-reviewed articles and records of controlled trials. Reference sources including books or book chapters, conference publications, commentaries, editorials and grey literature were excluded. A systematic search was performed on the following databases: i) PubMed ii) Embase and iii) the Cochrane Central Register of Controlled Trials (CENTRAL), using the search criteria summarized in Table 1.

**Table 1:** Database search strategy, search terms and filter criteria.

| Database | No. | Search term | Study type | Publication date | Raw hits | Filtered hits |
| --- | --- | --- | --- | --- | --- | --- |
| **PubMed**<br>**Search date:**<br>**2024/05/07**<br>**Repeated search date:**<br>**2024/05/10** | #1 | ((malaria[Title/Abstract]) AND (pregnancy[Title/Abstract]) AND ((inflam*[Title/Abstract]) OR (angiogenesis[Title/Abstract]))) ) AND (birth[Title/Abstract]) | 1. Clinical Trial<br>2. Meta-Analysis<br>3. Randomized Controlled Trial | 2004/1/1 to 2024/5/1 | 100 | 7 |
| **PubMed**<br>**Search date:**<br>**2024/05/07**<br>**Repeated search date:**<br>**2024/05/10** | #2 | ((pregnancy[Title/Abstract]) AND (inflame*[Title/Abstract] OR angiogenesis[Title/Abstract])) AND (birthweight[Title/Abstract] OR birth outcome[Title/Abstract]) |  |  | 759 | 47 |
| **Embase**<br>**Search date:**<br>**2024/05/07**<br>**Repeated search date:**<br>**2024/05/10** | #1 | (('malaria'/exp OR malaria) AND pregnancy:ti,ab,kw AND inflammation:ti,ab,kw OR 'angiogenesis modulator':ti,ab,kw) AND 'birthweight':ti,ab,kw AND [2004-2024]/py | 1. animal experiment<br>2. animal model<br>3. cohort analysis | 2004 to 2024 | 500 | 20 |
| **Embase**<br>**Search date:**<br>**2024/05/07**<br>**Repeated**<br>**search date:**<br>**2024/05/10** | #2 | #1 AND ('animal experiment'/de OR 'animal model'/de OR 'cohort analysis'/de OR 'controlled study'/de OR 'ex vivo study'/de OR 'in vitro study'/de OR 'in vivo study'/de OR 'major clinical study'/de OR 'mouse model'/de OR 'multicenter study'/de OR 'observational study'/de OR 'prospective study'/de OR 'randomized controlled trial'/de) | 4. randomized controlled trial<br>5. ex vivo study<br>6. in vitro study<br>7. in vivo study<br>8. clinical study<br>9. mouse model<br>10. multicenter study<br>11. observational study<br>12. prospective study |  |  |  |
| **Cochrane (CENTRAL)**<br>**Search date:**<br>**2024/05/07**<br>**Repeated**<br>**search date:**<br>**2024/05/10** | #1 | ("malaria control intervention"):ti,ab,kw AND ("pregnancy"):ti,ab,kw AND ("inflammation"):ti,ab,kw OR ("angiogenesis"):ti,ab,kw AND ("birthweight"):ti,ab,kw (Word variations have been searched) | Trials:<br>1. PubMed (30)<br>2. Embase (26)<br>3. CT.gov (5)<br>4. ICTRP (1) | 2004 to 2024 | 41 | 40 |

### Database search strategy

The search terms “pregnancy”, with or without “malaria”, and “inflammation”, “angiogenesis”, “birthweight”, or “birth outcome” were used to review likely interplay of inflammation and/or angiogenesis on infant birthweight. Drug names (sulfadoxine-pyrimethamine) were purposefully not included to expand the search beyond malaria-specific studies and to capture studies describing non-malarial pathways that may contribute to birthweight outcomes. The search strings were adapted to the functionalities and features of the databases (summarized in Table 1) and results were filtered based on included study types, date of publication, and language (English). This initial database search was conducted by the first reviewer (SS) on 07 May 2024. Additionally, we examined the reference list of included articles and tracked any subsequent publications from relevant clinical trials reported after the search date.

### Study selection: Screening and eligibility criteria

All downloaded citations were imported to reference managing software Endnote before exporting them to an online systematic review software, Rayyan[29]. The study selection was conducted within the Rayyan interface, and the workflow was recorded using a PRISMA flowchart. The first reviewer (SS) drafted the list of eligibility criteria for data screening, which was adapted and finalized in consultation with the second reviewer (PM) for improving clarity and reproducibility (summarized in Table 2).

**Table 2:** Eligibility criteria for data screening.

| No. | Search criteria | Description (comma separated indicates OR) |
| --- | --- | --- |
| 1 | Study type | Randomized controlled trials, observational studies, association study, cohort analysis, ex-vivo, in-vivo, in-vitro animal studies (mouse model), systematic review & meta-analysis |
| 2 | Publication type | Peer-reviewed, recorded in clinical trial registry (CT.reg, ClinicalTrials.gov) |
| 3 | Language | English |
| 4 | Population | HIV-uninfected pregnant women: healthy obstetrical patients, or patients with *Plasmodium falciparum* malaria infection (with or without co-infection), or systemic inflammation (periodontal infection [30] and/or inflammatory bowel disease [31])(commonly recorded during pregnancy and women in reproductive age); Animal models: pregnant mice infected with rodent-specific *Plasmodium* strains (such as *P. berghei*, *P. chabaudi*) |
| 5 | Intervention (for controlled trials) | Antimalarial drugs indicated for IPTp use (such as, sulfadoxine-pyrimethamine)<br>Treatment of systemic inflammation such as periodontal treatment |
| 6 | Mediator | Inflammatory and/or angiogenesis biomarker |
| 7 | Outcome | Infant (neonatal) birthweight |

As this review aimed to examine non-malaria attributable risk of low birthweight from mothers who had spontaneous pregnancies, we excluded studies that focused on related but secondary outcomes (such as, pre-term birth or neo-natal survival, or both), and/or assessed the effect of other intervention (such as, in-vitro fertilization, cholecalciferol supplementation etc.). Additionally, we excluded patients with HIV co-infection, since IPTp-SP is contraindicated in this sub-group[5].

Titles and abstracts of all articles were first screened by both the first and second reviewers independently with “blind on” mode in Rayyan software based on these inclusion criteria. A high convergence rate between reviewers was achieved (95.6%) for included articles. The two reviews resulted with conflicting assessments for four articles. Full text versions of these were reviewed independently by both reviewers. This led to the inclusion of an additional article, and rejection of three.

### Data extraction

The detailed description of the charted data items and charting process is presented in the supplementary materials (supplementary section 1.1, Table S1, section 1.2). We extracted data from the included studies in a table format containing relevant details: study setting (country), year of publication, study type, cohort characteristics, primary outcomes, intervention (if any), key results, data access information, reference including the first author’s name and citation. No critical appraisal of individual sources of evidence was conducted in the scope of our study. However, only peer-reviewed studies from relevant databases were included for data extraction and result synthesis (supplementary Table S2).

The included study results were categorized based on the study type: i) randomized controlled trials (RCT) or prospective cohort studies including participants with malaria infection or conducted in malaria endemic settings ii) RCT or prospective cohort studies or observational studies without having malaria infection iii) systematic reviews and meta-analyses and iv) pre-clinical studies using rodent models.

### Synthesis of results

We reviewed the full text of all the included studies to gather information on the study setting, methods used and relevant results. First, we collated general characteristics of the study (such as, settings and designs). Next, the list of primary study outcomes including time point(s) of data collection were collected. Key findings from the included studies were examined to synthesize our results on: i) which inflammatory and/or angiogenesis regulators likely influenced the low birthweight risk based on the likely association as reported in the study (such as, adjusted odds ratios from the regression analysis) and ii) potential impact of IPTp drugs on these biomarkers. These results were broadly categorized by the study types. A conceptual causal diagram was constructed for visualization and communication of the likely pathways involved and putative interactions with IPTp drugs. Finally, on the basis of this investigation, some data needs were identified.

## Results

### Search results

We identified a total of 1773 articles from a search of three databases. After removing non-relevant studies (n=1638) based on the pre-set filter criteria and duplicates (n=24), a total of 111 articles were examined. Two additional studies were detected through citation searches[32, 33], while one further study was obtained from a website search to follow-up on a recently concluded trial that was published after the date of our database search[22]. The complete process of examination and inclusion is outlined in Fig. 1. Altogether, a total of 15 eligible studies were included for data extraction and charting (supplementary Table S2).

**Fig. 1:**
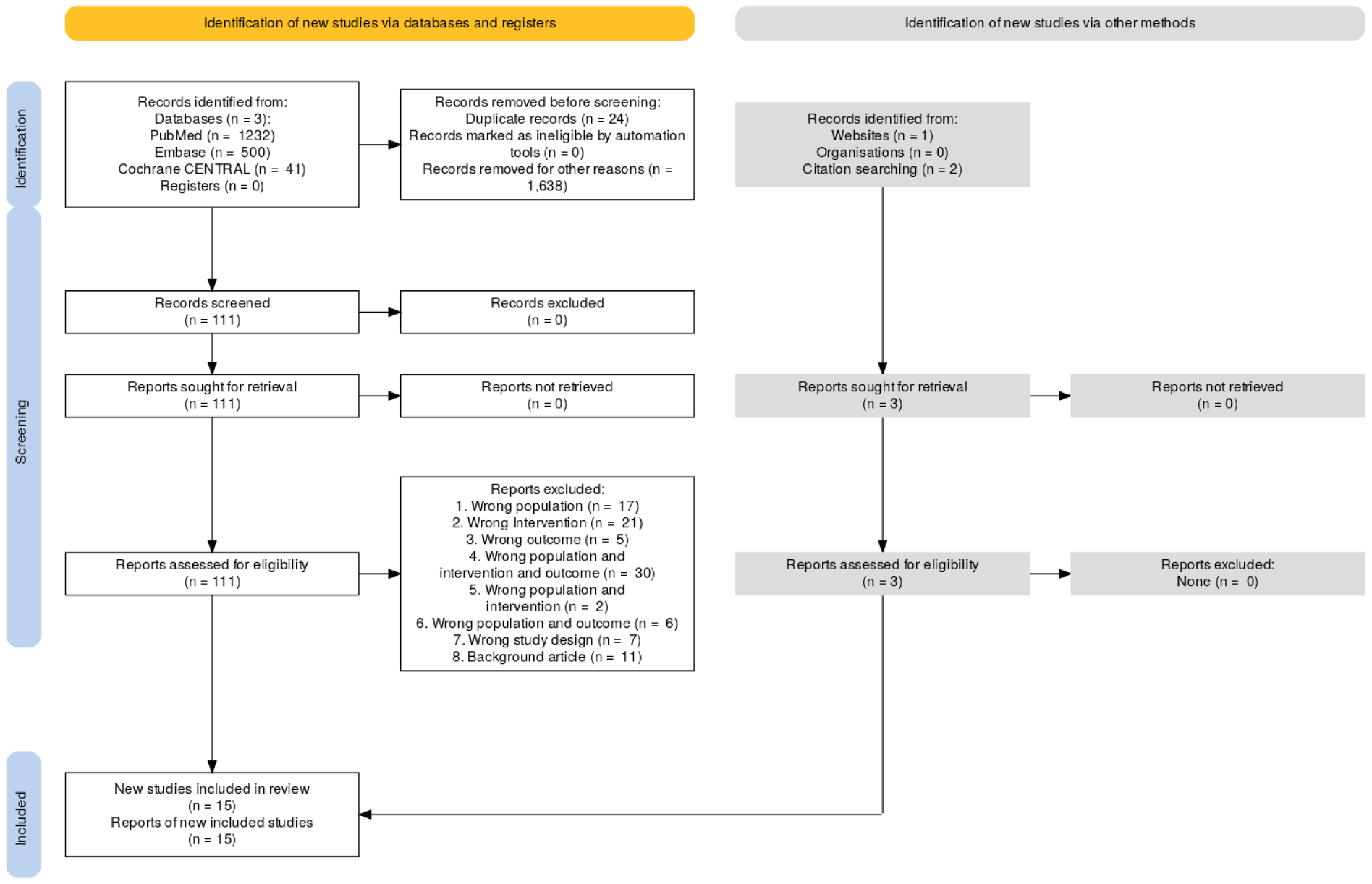
PRISMA flowchart depicting the retrieval and screening of peer-reviewed publications. For each database screened, publications were screened for exclusion criteria by automatic filter to identify eligible study types (i.e. controlled trial, systematic review, meta-analysis and pre-clinical study). Exclusion criteria are as follows: 1) *wrong* population (e.g., pre-mature infant with retinopathy, pregnant women having in-vitro fertilization) 2) *wrong* intervention (e.g., dietary supplement) 3) *wrong* outcome (e.g., pre-term birth or maternal health outcome) 4) a combination of *wrong* population and *wrong* intervention and wrong outcome 5) a combination of *wrong* population and intervention 6) a combination of *wrong* population and outcome 7) *wrong* study design (e.g., biomarker data not recorded) 8) background article (informative but without direct association to this review). Note: ‘*wrong*’ simply indicates ‘other’ than relevant in the context of this review. Three additional studies were included from PubMed through citation and website searches.

### Study settings and designs

Of the studies included in this scoping review, 8 out of 15 (53%, value rounded to whole number) involved pregnant women with malaria infections and living in malaria endemic regions[19, 22, 34–39]. while 4 out of 15 (27%) involved healthy pregnant women or women with periodontal diseases but without malaria[30, 32, 33, 40]. Only two clinical trials (13% of all studies) examined IPTp-SP compared to other alternative chemoprevention drugs (chloroquine[19] and DP[22]), two studies (13%) were interventions against pregnancy specific periodontitis [40] or severe gingivitis [30] while all others were prospective cohort studies or observational studies of longitudinal birth cohort data. The majority of RCTs conducted on women with malaria or living in malaria endemic areas were conducted in Africa (6/8), while longitudinal analyses of birth cohort data were reported from island countries in east Africa (Seychelles) and Southeast Asia (Philippines). Studies investigating periodontics were conducted in the USA and UK (Fig. 2). Additionally, 1 out of the 15 (7%) publications was a systematic literature review that incorporated most of the studies conducted in Europe and North America[41] and 2 out of 15 (13%) were pre-clinical genomics studies conducted on mouse models [23, 42]. In one of these, pregnant mice were infected with *P. berghei,* a *Plasmodium* species that infects non-human hosts.

**Fig. 2:**
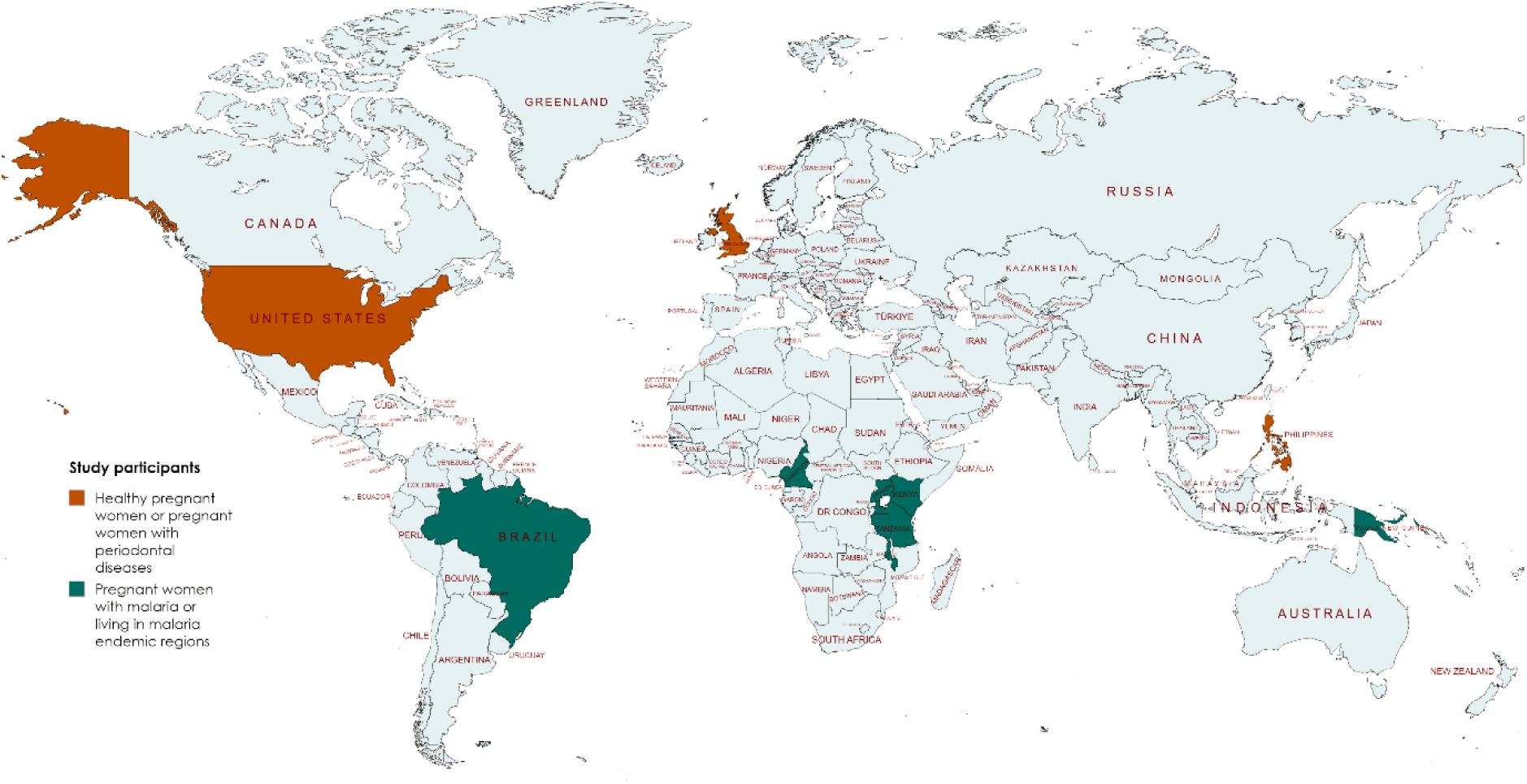
Geographical locations and cohort characteristics of randomized controlled trials and association studies included in this investigation. The studies examining pregnant women with malaria or living in malaria endemic regions are marked with green, and studies examining healthy pregnant women or with periodontal infection are marked with dark orange. Systematic reviews, and pre-clinical studies conducted on rodent models are not delineated in this figure. The world map is coloured as per study cohort type using the online tool www.mapchart.net.

### Outcome measures

The primary outcome of the included studies varied substantially, based on the study aims for each. This included measures of: **1) markers of inflammation (including both pro- and counter-regulatory molecules)**: i) C-reactive protein (CRP) [19, 22, 32, 33, 40] ii) α-1-acid glycoprotein (AGP) [19, 22] iii) soluble endoglin (sEng)[19], iv) tumor necrosis factor alpha (TNF-α) [19, 30, 32, 33, 37, 38, 40] v) anti-*Pf*GPI antibodies (immunoglobulin M – IgM and IgG levels)[43] vi) soluble TNF-αR (sTNFR) 1 and sTNFR2[39] vii) markers of IFN pathway (IFN-β[36], IFN-γ [33, 37], IL-10 [32, 33, 36–38], IL-β [30, 33, 40], IL-6 [30, 32, 33, 40]and MX-1[36] vii) Prostaglandin E2 (PGE2)[40] viii) CCR5[42] ix) MyD88[44] 2**) markers of placental angiogenesis**: i) soluble fms-like tyrosine kinase- 1 (sFlt-1) [19, 33] ii) placental growth factor (PlGF) [19] iii) sFlt-1/PlGF ratio[19] iv) markers of complement activation (such as, C3a, C4a, C5a) [38] v) vascular endothelial growth factor (VEGF)- D[33] **3) markers of neutrophil activation**: i) myeloperoxidase (MPO) [34] ii) proteinase 3 (PRTN3) [34]iii) matrix metalloproteinase (MMP9) [34, 40] **4) infant health outcomes**: i) low birthweight ii) preterm birth[19, 22, 36, 40, 42, 43] iii) small-for-gestational age [19, 22, 34, 36], iv) fetal loss[22] and neonatal death[22]. One study recorded the relationship of ferritin and leptin levels in the cord blood to birthweight[37]. Additionally, parasitaemia was assessed in studies that included participants with malaria infection, or living in malaria endemic region [34, 43].

### Data collection and access

Both maternal biomarker levels and infant anthropometric measures were recorded at delivery in the majority of studies that included pregnant women with malaria or conducted in malaria endemic regions[19, 34, 36, 38, 39, 43]. Two recent clinical trials also collected baseline inflammatory or angiogenic biomarker data at enrolment[19, 22] and another observational study collected blood smear samples to detect malarial infection after any fever episode[36]. We found only one study where such data were collected at multiple time points throughout pregnancy (at every monthly visit after 13th weeks of gestation) and at delivery[39]. The primary outcomes were segregated by gravidity in some association studies [34, 36, 38, 39, 43], but not in clinical trials that investigated the impact of IPTp on inflammatory or angiogenic biomarkers.

Among clinical studies or association studies incorporating healthy pregnant women or with periodontal infections, the biomarker data were collected at variable time points based on study aims: between 13- 16 weeks and 29-32 weeks of gestation[40], 16 to 24 weeks of gestation[30]. 28 weeks of gestation[33] or third trimester[32] and at delivery[30, 40].

Individual level data from two trials are available from the World Wide Antimalarial Research Network (WWARN), that require access approval by the WWARN IDDO Data Access Committee[19, 22] and in PRIDE repository (https://www.ebi.ac.uk/pride)[38], respectively. Two studies reported aggregated data that are noted as available upon author request [34, 44], while data sharing information was not indicated in the majority (60.00%, n=9) of the included studies.

### Study findings leading to result synthesis

The data charted from the literature review indicated that inflammation or dysregulation of angiogenesis, or both, could be significant determinants of low birthweight delivery. Several studies that lacked mediator biomarker records were excluded despite their investigations of the associations between LBW with chronic inflammatory diseases or systemic inflammation. Insights from the articles reviewed as part of this study are described in detail in Supplementary Table S2.

## 1. Impact on inflammatory biomarkers

### 1.1 Results from RCTs or prospective cohort studies incorporating women with malaria infection or living in malaria endemic regions

Data from two clinical trials were identified which explicitly investigated the impacts of IPTp drugs on inflammatory biomarkers[19, 22]. In these studies, the levels of CRP and AGP were measured in maternal venous plasma samples at enrolment and at delivery. A significant association of low CRP level with improved birthweight was observed in one of these trials conducted in a low malaria prevalence area of Papua New Guinea (PNG)[19], but such an association was not detected in the other trial conducted in a high malaria prevalence setting in Malawi [22].The Malawi trial [22]found that the women with higher AGP levels at delivery and who received DP or DP+AZ had higher risk for adverse birth outcomes including low birthweight, pre-term delivery and foetal loss (adjusted rate ratio 1.6), while women who received SP did not show this association. Also, women receiving SP+AZ had lower AGP levels and improved birthweights in the PNG trial[19].

The potential association of inflammatory biomarkers to birth outcomes was explored from four cohort studies [36–39]. Higher levels of some pro-inflammatory biomarker in placental blood reflective of systemic inflammation (such as Type II interferon – IFN-γ) have been correlated with lower birth weight was reported [37]. Given the levels of IFN-γ in the peripheral blood were not increased while the same increased substantially in the placenta, the authors of that study speculated that both placental and foetal cells were the likely sources for the production of local inflammatory markers. In another cohort study, the levels of soluble TNFR1 and TNFR2 (counter-regulator of TNF-α pathway) were negatively associated with birthweight[39]. The dynamic changes in sTNFR levels were monitored longitudinally at monthly time intervals. However, the aim of this study was to examine the value of sTNFR as a diagnostic biomarker of placental malaria and its potential association with infant health outcomes. Thus, data on the antimalarial dosing regimens and any impact on these biomarkers was not recorded. In another study, it was reported that the levels of IL10 were higher both in human and in mouse models infected with *P. falciparum,* and *P. berghei*[38], respectively, but such an effect on foetal growth was not clear.

Elevated immunity markers of interferon type I pathway, particularly IFN-β levels, were found to be protective against foetal growth restriction in a follow-up genomics analysis[36] utilizing data from a RCT conducted in high malaria prevalent settings in Uganda[45]. Higher IFN-β levels were correlated with improved birthweight, which increased with gravidity. Although the initial clinical study investigated IPTp-SP vs. DP impact on malarial outcomes, the follow-up study investigated the association of IFN-β with infant health outcomes based on gravidity. However, these secondary outcomes were not reported by trial arms. This limited our ability to draw conclusions on SP vs. DP’s effect on this protective immunity directed pathway. Similarly, some markers in the cord blood (such as, ferritin) were positively associated with birthweight in another study but the impact of chemoprevention drugs on these were not evaluated [37].

### 1.2 Results from RCTs, prospective cohorts or observational studies incorporating healthy pregnant women or women with periodontal diseases

In two birth cohort analyses of infants born from healthy pregnant women, the ratio of pro- to anti-inflammatory markers (such as, IL6/IL10 ratio) and CRP (as a composite measure of inflammatory processes) in the mother was associated with infant birthweight. Interestingly, there was no such association when each cytokine alone was evaluated [32, 33]. Another association study investigating the impact of non-surgical therapy on pregnant women with periodontitis found no correlation of serum inflammatory biomarkers tested (such as, CRP, TNF- α) at 13-17 weeks or 29-32 weeks of gestation to birth weight[40]. Notably, this study excluded women who required any antibiotic treatment during pregnancy. The authors attributed the lack of association to the absence of antimicrobial agents and the small sample size.

### 1.3 Results from animal experiments

Elevated levels of the chemokine receptor CCR5, a factor which is known to regulate immune response during inflammation, were associated with lower birth weights in a preclinical model in which mice were infected with *P. berghei*[42]. Another genotyping study examined the interplay of the potentially variable influence of maternal and foetal immune response on offspring anthropomorphic measures. This was estimated by quantifying the expression levels of MyD88, an adapter protein involved in regulating the production of pro-inflammatory proteins. Interestingly, only the levels of foetal-derived MyD88 were associated with placental malaria and low birth weight, which suggested a mechanism of action involving the foetal triggering of inflammatory pathways[44].

## 2. Impact on angiogenic biomarkers

Malaria in pregnancy may lead to an imbalance in levels of angiogenic factors (such as, VEGF, sFlt1, soluble endoglin, PIGF) involved in placental vasculogenesis. Elevated markers of the complement activation pathway were found to increase the risk of angiogenesis dysregulation, placental inflammation, intrauterine growth retardation, low birth weight and preterm delivery[19, 33]. A clinical trial investigating the impact of IPTp with SP+AZ on infant health outcome revealed dysregulation of placental angiogenesis factors; there, an increased sFlt-1/PlGF ratio at enrolment was associated with low birthweight delivery in women with or without malaria. This indicated the likely involvement of this pathway regardless of infection type[19]. Consequently, low PlGF levels and high sFlt-1:PlGF ratios are suggested to be useful predictors of adverse pregnancy outcomes, particularly in the third trimester of pregnancy. One study analysed proteomic markers and found a negative correlation between complement system activation markers, such as complement C1r subcomponent-like protein, C4-A, and birthweight in pregnant women infected with *P. falciparum,* and mouse model infected with *P. berghei*[38]. Notably, in a study by Yeates and colleagues, birth cohort analysis of infants born from healthy pregnant women indicated that angiogenesis dysregulation even without specific infection may lead to LBW delivery, observed as elevated VEGF-D in later pregnancy, when the higher level is typically undesirable[33].

## 3. Impact on neutrophil activation

Granulocytes have been found to be relevant to malaria in pregnancy. Notably, the regulation of granulocytes is likely different in peripheral vs. placental blood based on placental malaria. Although neutrophil activation markers were found to be elevated in the placenta of a pregnant women diagnosed with malaria, their impact on either maternal or foetal health has so far not been well understood[34]. Further investigations would help us to better understand this pathway including any interaction with antimalarial drugs.

## 4. Potential non-malarial effects of IPTp-SP

Based on our investigation, we hypothesize that multiple underlying non-malarial effects of IPTp-SP may contribute to its effect on women and infants in the context of malaria in pregnancy, including: i) anti-inflammatory effect[16, 17, 19, 46] ii) prevention of pathological angiogenetic dysregulation[19, 33, 38] and iii) increasing maternal gestational weight (likely due to preventing inflammation and supporting shift towards healthier microbiota balance)[19]. Our observations are consistent with findings from a previous systematic review by Gomes and colleagues that examined the levels of maternal blood biomarkers of inflammation, growth and hormonal profile to birth outcome[41].There, the authors reported that increased levels of pro-inflammatory markers (such as, CRP, TNF-α, IL-ß, IL- 6) and upregulated hormonal biomarkers (such as, cortisol, ß-hCG) were likely to be associated with oxidative stress, preeclampsia and dysregulation of placental vasculature contributing to LBW. The study by Gomes and colleagues did not review the potential impact of any drug, and we also found only two studies that explicitly investigated the IPTp impact on biomarkers[19, 22]. However, we considered a potential interaction between the putative pathways for maternal inflammation and for the presence of angiogenic biomarkers and how this relates to birthweight of neonates and infants. In this scenario, we speculate on the potential impact of IPTp on such pathways, whereby a variety of non-malarial effects of SP (if any) might contribute to variability in birthweight and other infant health outcomes (**Fig. 3**). As shown, the scenarios are testable for each treatment (or drug) individually or as combination when given for IPTp use case.

**Fig. 3:**
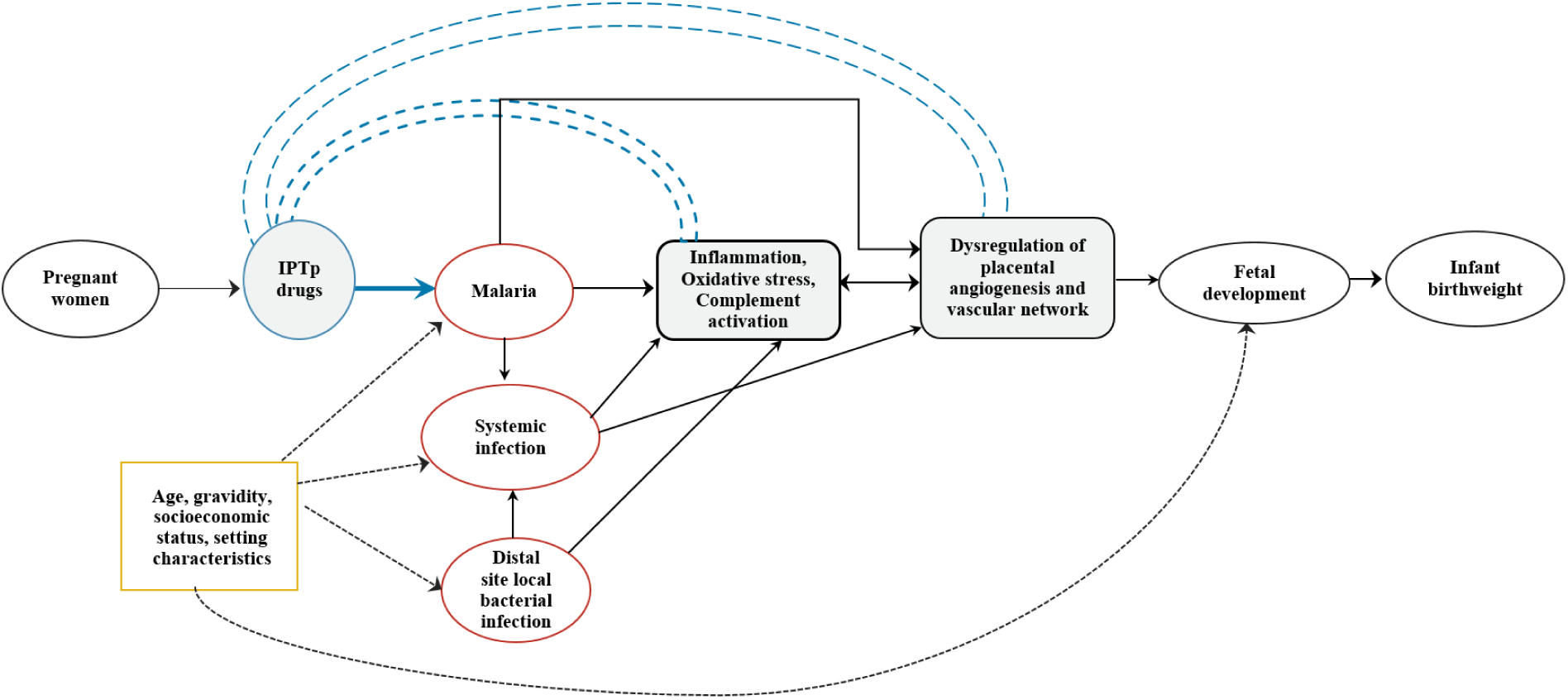
Conceptual causal diagram of potential non-malarial effects of antimalarial drugs on birthweight. While this figure is not based on formal causal effect analysis, it visually represents the results from included studies. Each component of this diagram represents generic pathways likely involved in maternal to foetal and infant health outcomes (birth weight). Exposure, intermediate state, and outcome variables are depicted as white ovals with black borders. Potential disease states, such as malaria and other infections, are shown as white ovals with orange borders. Background risk factors are represented by white boxes with yellow borders. The intervention (IPTp) is depicted as a grey circle with a blue border. Likely pathophysiological factors influencing birthweight are shown as grey squares with black borders. Black solid arrows represent relationship of disease states contributing to the outcome variable. Black dotted lines indicate the potential effects of background risk factors on disease states and intermediate variables. Potential non-malarial effects of IPTp drugs (e.g., IPTp-SP) are shown by blue dashed double lines. IPTp: intermittent preventive treatment in pregnant women; SP: sulfadoxine- pyrimethamine.

## 5. Gaps in current studies, and proposed action measures for future studies

Presently, it is understood that the total favourable impact of IPTp drugs (SP) especially on infant health outcomes may include both antimalarial and non-malarial effects[19]. But we so far do not fully understand the downstream effects and timeframes for such phenomenon, including how pro- and anti-inflammatory or angiogenesis mediators might be relevant throughout the gestation timeline. In an attempt to address the putative gaps in our knowledge and evidence related to the non-malarial benefits of IPTp treatments on maternal to infant health, we propose the following action and outcome measures to consider for future studies in this area of investigation (**Table 3**).

**Table 3:**
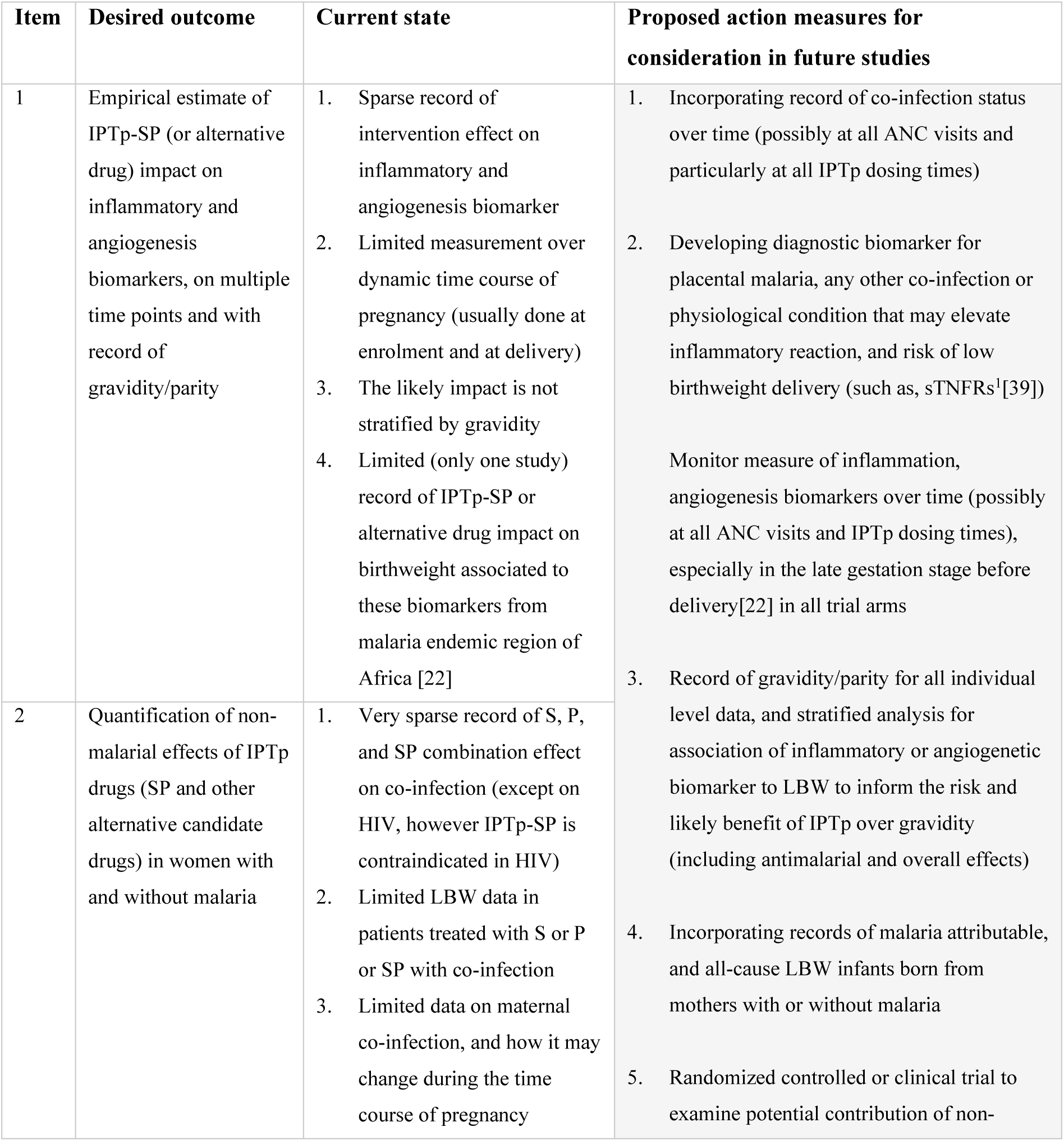
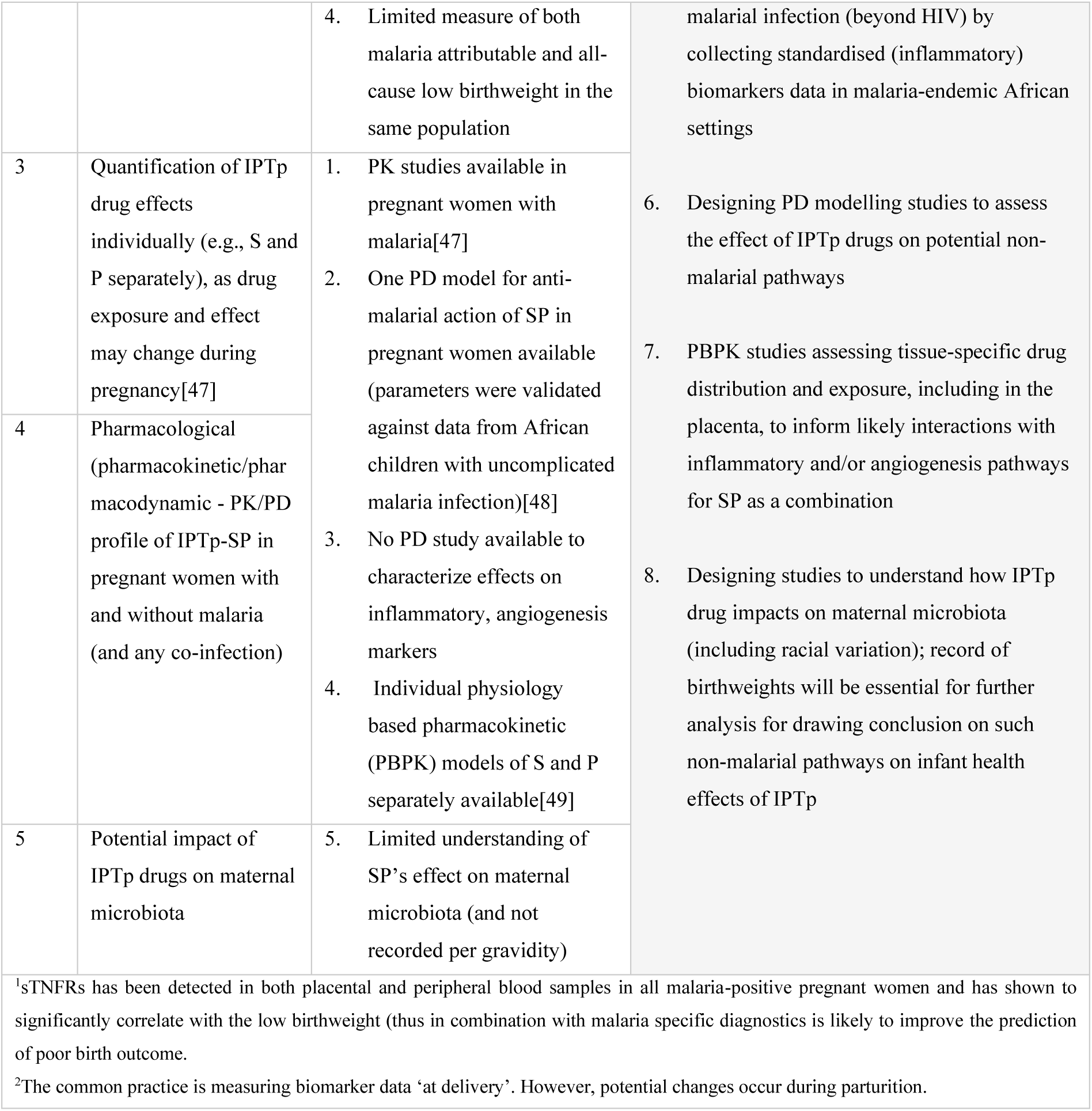
Proposed considerations for study designs and outcome measures.

## Discussion

In this scoping review, we synthesised new insights regarding putative non-malarial benefits of IPTp-SP treatments by analysing published articles derived through specific keyword searches of PubMed, Embase and Cochrane CENTRAL databases. This systematic scoping of literature enabled us to examine study methods including reported measures of secondary endpoints and other relevant information across a broad range of empirical studies. We summarised current knowledge and identified knowledge gaps in this area of research[27]. We endeavoured to propose potential action measures to address such gaps in future studies. The findings from this study are beneficial in the following three ways. First, comprehensively define the characteristics of the current standard-of-care SP as a comparator for alternative IPTp candidates. Second, evaluate the contribution of non-malarial risks to total LBW risk over time, thereby informing optimisation of timing and deployment strategies of IPTp and other non-malaria directed treatment [5]. Third, improve understanding of why SP maintains effectiveness in partially resistant settings[5, 7, 50]. Given that SP is widely used across the most vulnerable populations (both children and pregnant women[5]), a more complete understanding of its effects is crucial.

Some recent trials revealed a greater impact of IPTp-SP on birthweight compared to antimalarial DP, the latter of which is deemed more efficient as a treatment drug[19, 22]. Thus, SP has been postulated to benefit infant health outcomes beyond its antimalarial effect[19, 51], even beyond its broad spectrum antibiotic effects. However, the current evidence for the putative impacts of SP on bacterial infections during pregnancy and its influence on foetal growth is inconsistent. Two clinical trials reported an effect of IPTp-SP against only one bacterial infection *(Chlamydia Trichomonas)*[16, 17]. Surprisingly, this effect was greater compared to IPTp with anti-chlamydial drug AZ. Since the effects of SP on other microbial infections were comparatively inferior, we surmise that it would be informative to assess the potential antibiotic effects of sulfadoxine and pyrimethamine individually and as a combination by dedicated microbiological assays on samples collected from pregnant women with or without malaria in pregnancy (MiP).

Recent trial results indicated added benefits that may stem from the impact of IPTp-SP on chronic inflammation at sites local or even distal to the placenta (e.g., periodontitis), in intriguing[30, 40, 41] . Since the levels of inflammatory biomarkers measured in an individual can be influenced by both *Plasmodium* and non-malarial (such as *Chlamydia*) infections, preventing both such infections would increase the benefits of this treatment to limit LBW[19]. Some cohort studies conducted on women with malaria or other distal site infections (e.g., periodontitis, gingivitis)[30] showed positive correlations between reduced inflammation marker levels and increased birthweights. Notably, the balance of pro- to anti-inflammatory markers (such as IL6/IL10 ratio or CRP as a composite measure) is more likely to be found to be significant when associated with infant birth outcomes rather than any such individual biomarker[32]. However, none of these clinical studies included SP. Therefore, we conclude in this study that a dedicated measure of the impact of SP impact on a broad range of inflammatory markers, including markers that are representative of the balance between pro- to anti-inflammatory processes, is needed. Moreover, because gravidity is likely a confounder in statistical analyses and IPTp is recommended based on gravidity it will be useful to record gravidity information.

Any anti-inflammatory benefit would be beneficial to improve the gestational weight of the mother and, in turn, serve as a predictor of healthier infant weights[18]. SP has been suggested to influence gut microbiota composition and energy balance, however, there is currently little understanding of the impact of SP on birthweight in terms of possible interactions with the maternal microbiota. Results from a recent controlled trial revealed that the benefit to be attributable to the antibacterial properties of SP and its protective effect against common intestinal pathogens (such as, atypical enteropathogenic *E.coli*/enterohaemorrhagic *E. coli* and typical enteropathogenic E.coli)[51]. The lower enteric infection burden in mothers in the SP arm of that trial was correlated with higher gestational weight gain and improved birthweight compared to DP. This finding is consistent with findings from a recent trial which found that the impact on placental and maternal clinical malaria remained comparable between the SP and DP groups, however the positive effect on LBW was higher in the SP group (LBW 7.2% vs 10.3% in DP group)[16]. Also, genetic heterogeneity of the participants in the study would have further accounted for variability maternal microbiome (e.g., vaginal microbiota)[52]. The precise pathways that underlie the mechanistic impacts of SP and its related treatments need further exploration, including the influence of host genetics and other demographic variabilities.

A unique pregnancy specific pathway by which SP may influence the birthweight includes the regulation of placental vasculogenesis which, in turn, could directly contribute to nutrient and hormonal exchange that supports robust foetal growth. For instance, low levels of pro-angiogenic PIGF, high levels of anti-angiogenic sFLT1 and a high sFLT1/PIGF ratio (commonly also used for diagnosis of pre-eclampsia) are found to possibly predict the risk of adverse pregnancy outcomes including LBW and small for gestational age[19]. The impact of SP on angiogenic markers is likely to be important for predicting its total effect[19, 32, 41]. Some pathological angiogenic dysregulations might be preventable by SP (with AZ)[19]. However, the individual impact of SP on these parameters is currently not known, nor how it changes with gravidity. The authors noted that results from a new trial[22] may further expand our understanding of the impact of SP on relevant biomarkers. Encouragingly, the trial[22] focused on anti-inflammatory markers, however the effect on angiogenesis remained uncharacterized.

Our study recognises the importance and potential benefit for estimating LBW risk that is attributable to both malaria and non-malaria effects according to gravidity, for different IPTp dosing schedules[5]. Currently, there is very a limited understanding of the differential non-malarial impact of IPTp drugs by gravidity. This is partly because gravidity has been considered a confounder in statistical models from trials. As a result, only single estimates for the drug impact across gravidity levels are reported in existing trials. For instance, the IMPROVE trial addressed many other data gaps including collecting gravidity information[22]. However, a single estimate of association was reported for all pregnant women. It will be of interest to estimate such association per gravidity in future trials.

Maternal physiology, including changes in biomarker levels, change dynamically throughout pregnancy. For instance, VEGF-D is a placental angiogenesis marker and elevated levels in late pregnancy has been shown to be negatively associated with birthweight, while a positive association is observed during early pregnancy. This is supposedly due to the dynamic changes required for VEGF- D levels for the normal course from early to late pregnancy[33]. Thus, it will be useful to track marker levels at multiple time points throughout the pregnancy to predict the drug effect based on dosing schedules[22, 33, 41].

With drug treatments, knowledge of the pharmacokinetic and pharmacodynamic (PK/PD) characteristics are crucial for estimating intervention effect in heterogeneous populations[53]. For example, altered pharmacokinetics during pregnancy can explain increased clearance of sulfadoxine by three-fold (subsequently decreased during post-partum period), while the clearance of pyrimethamine was reduced by about 18%[47]. Thus, the potential anti-inflammatory effects of SP during pregnancy is likely contributed by relatively higher pyrimethamine concentrations. However, there is currently insufficient data to conclude individual effects of individual drugs on biomarkers for inflammation and angiogenesis. While the anti-inflammatory effects of sulfadoxine have been assessed to some extent, investigations on such effects with pyrimethamine treatment have only recently begun to be explored[23]. Further research is needed for individual and combination drugs for characterising the non-malarial PK/PD effects, based on data from subjects with or without malaria, and from control groups. Additionally, physiologically based pharmacokinetic (PBPK) models that consider variation in drug disposition and metabolism, based on physiological difference between patients (among other system parameters), could be useful for understanding the drug disposition in different tissues to identify likely sites of off-target drug interaction[49]. Notably, while PBPK models are often developed for predicting drug-drug-interaction or dose adjustment based on pharmacokinetic properties in special population (such as, pregnant women), it will be beneficial to use such detailed PK information to further explore the exposure dynamics and the effect of pregnancy on drug PK profile.

One finding in our study was that, the levels of some inflammatory markers change only in placental blood but not in peripheral blood (such as, IFN-γ[37]). It is found that either or both maternal and foetal cells present in the placenta may express these biomarkers[37]. Therefore, identifying and validating the specific diagnostic sample type will be beneficial for predicting the effects of drug treatment [33, 37]. The concentrations of antimalarial drugs in different organs in pregnant women likely derived from PBPK modelling studies as previously described, might be additionally informative[49].

This study included a range of study types, spanning randomized controlled trial, prospective and birth cohort analysis, meta-analysis in pregnant women with and without malaria and animal studies to enable synthesizing collated insights from a variety of perspectives. Using this rich source of information, we explored both the knowledge gaps and proposed potential outcome measures that can be considered for expanding data quality and types. Second, we included other common infections during pregnancy (particularly periodontal diseases) to examine the association of inflammatory and/or angiogenic biomarkers with infant health outcomes independent of malaria. Third, the included studies were conducted in varying geographical locations. While this was not a selection criterion, it may help to reduce the influence of external factors on biological processes (such as, access to treatment due to socio-economic differences), and contribute to expanding our understanding on additional relevant factors (such as differential microbiota linked with racial variability). Finally, although this study focuses on pregnancy outcomes in humans, we included relevant studies on animal models to further advance the understanding of complex pathological processes that may not have been studied in pregnant women. Findings from these studies are likely to generate evidence on the contribution of maternal malaria to other coinfection (such as, the elevated levels of CCR5 in a study with mice model indicated higher risk of HIV transmission from mother to foetus, similar to what is proposed for pregnant women affected with *P. falciparum*)[42]. Some studies found consistent results from human and mouse pregnancies[38], although drawing direct conclusions is not always possible. Nevertheless, pre-clinical studies provide valuable insight to inform validation studies in humans[44]. Overall, our study considered multiple perspectives relevant for IPTp drug effects.

There are several limitations of the current study. First, we screened studies from three databases based on defined parameters and timelines which means that some studies may have been missed. Of note, additional searches beyond the defined search date for our database search led to the identification of several articles investigating the effects of IPTp-SP versus alternative drugs in the context of non-malarial effects[54, 55]. However, these were not included because each lacked biomarker data that were relevant to this study. Secondly, the choice of search terms was intended to capture both malaria-related, and other infections prevalent during pregnancy. As such, only periodontal infection was included as a relevant example of co-infection. Thirdly, studies published after the screening date, and studies published in languages other than English were not captured. Finally, we intentionally did not conduct any critical assessment of the reviewed studies, as it was more important for our scoping review to include versatile study types[26, 27] to examine pathways with which IPTp drugs may interact beyond malaria and clarify relevant concepts. These limitations are unlikely to change our broad conclusions.

## Conclusion

To better understand the overall benefits and effects of IPTp drugs in malaria, current evidence and data gaps call for future empirical studies to possibly record: i) maternal co-infections and microbiota information, ii) levels of inflammation and angiogenesis markers over time in all gravidities, and iii) information on both malaria attributable and non-malaria related low birthweight. It would be beneficial to examine whether gravidity is associated with the non-malarial effects of IPTp-SP, in addition to its antimalarial effectiveness in different drug sensitivity settings. Such comprehensive understanding may support decisions around IPTp strategies by taking into account both malaria- and non-malaria attributable risks and drug effects, to better align timing of interventions with these risks if informed by data across multiple gestational time points. Assessing the impact of IPTp drugs on non-malaria attributable low birthweight (and possibly other anthropomorphic measures, such as pre-term birth or small for gestational age) may support the design of preferred product characteristics of alternative drug candidates based on their total effect.

## Declarations

### Ethics approval and consent to participate

Not applicable. This study reported only publicly available anonymized data, no data was collected separately.

### Consent for publication

Not applicable.

### Availability of data and materials

All data analysed during this study are included in this article and its supplementary information file.

### Authors’ contributions

SS and MAP designed the study. SS developed and performed the database search. SS and PM conducted the screening, reviewing the articles. SS charted data and drafted the manuscript. PM, JJ and MAP validated the workflow, analyses, and results. All authors contributed to interpreting the results and making edits to the draft and final manuscript and gave their approval for publication.

## Data Availability

All data produced in the present work are contained in the manuscript and supplementary information.

## Acknowledgements

We acknowledge support from Julian Heng for English editing of the manuscript text. We appreciate all advice from the members of the Disease Modelling Research unit of the Swiss Tropical and Public Health Institute and thank Daria Hofer for the project management support.

## Authors’ information

**Swiss Tropical and Public Health Institute, Allschwil, Switzerland**

Swapnoleena Sen and Pablo Martinez de Salazar

**University of Basel, Basel, Switzerland**

Swapnoleena Sen, Pablo Martinez de Salazar and Joerg J Moehrle

**The Kids Research Institute Australia, Nedland, WA, Australia**

Melissa A Penny

**Centre for Child Health Research, The University of Western Australia, Crawley, WA, Australia**

Melissa A Penny

## Supplementary results

In this scoping review, we gathered evidence of the putative impacts of sulfadoxine-pyrimethamine (SP) beyond its antimalarial (anti-parasitic) effects when administered for intermittent preventive treatment. A systematic literature review of published randomized controlled trials, observational or pre-clinical studies, and meta-analysis studies was conducted on articles retrieved from PubMed, Embase and CENTRAL databases in May 2024. The screening, inclusion criteria and data extraction methods are detailed in the manuscript.

### 1.1 Data items

Here we first describe the data items in the context of our charted data, as per the essential checklists of PRISMA-ScR guidelines [1] (Table S1).

**Table S1:** List of charted data items from studies included in this investigation.

| Data Item | Description (including any assumptions and simplifications) |
| --- | --- |
| Setting | The country where the study was conducted (includes specific geographical location such as the city or province or state name if it was specified). This applies to randomized controlled trial (RCT) or association studies, while the broad range of regions or the relevant continent is indicated for systematic review and not applicable (NA) for animal models. Additionally, the study identifier is added for RCTs if reported within the study. |
| Study type | The methods applied in this study:<br>**i) RCTs** that included studies where randomization was applied on study cohorts.<br>**ii) Cohort study (prospective)** where no randomization was applied. The studies focused on examining the statistical association or correlation of study outcome to mediator investigated, but no intervention was tested.<br>**iii) Observational study (retrospective)** that included longitudinal birth data analysis.<br>**iii) Systematic review & meta-analysis** of publicly available articles and clinical trials data. |
|  | **iv) Animal study** where the study was conducted using only samples from animal models (such as, studies from mouse models). |
| Cohort | The study participants or study arms: i) healthy pregnant women ii) pregnant women with or without malaria ii) pregnant women with other infections (such as periodontal diseases) iv) pregnant mouse model |
| Outcome | List of primary outcomes as reported in the study. This included two types:<br>1) **Record of biomarkers measured:** i) inflammatory and/or ii) angiogenesis regulatory iii) mediator(s) of inflammatory and/or angiogenesis processes including counter-regulators, or genes involved in these processes<br>2) **Infant anthropomorphic measures** (including birthweight) |
| Data collection | Time points for study data (outcome measures) collected in the study |
| Intervention | Brief description of the intervention tested in the study (if any) |
| Study result | The key finding(s) from the study relevant to the objectives of our scoping review |
| Data access | Details about data sharing and access information as reported in the study (NR indicates this information was not reported) |
| Reference | Name of the first author, publication year and citation |

### 1.2 Data charting process

All charted data are summarized below, as per the listed data items from the included studies published between January 2004 to July 2024 (Table S2). Data charting was done independently by the first reviewer (SS), after the form (Table S2) was evaluated by both the first and second reviewer (SS, PM) for clarity of completeness of collated information. All reviewers re-checked the charted data tables after completion and the table was updated as necessary.

**Table S2:**
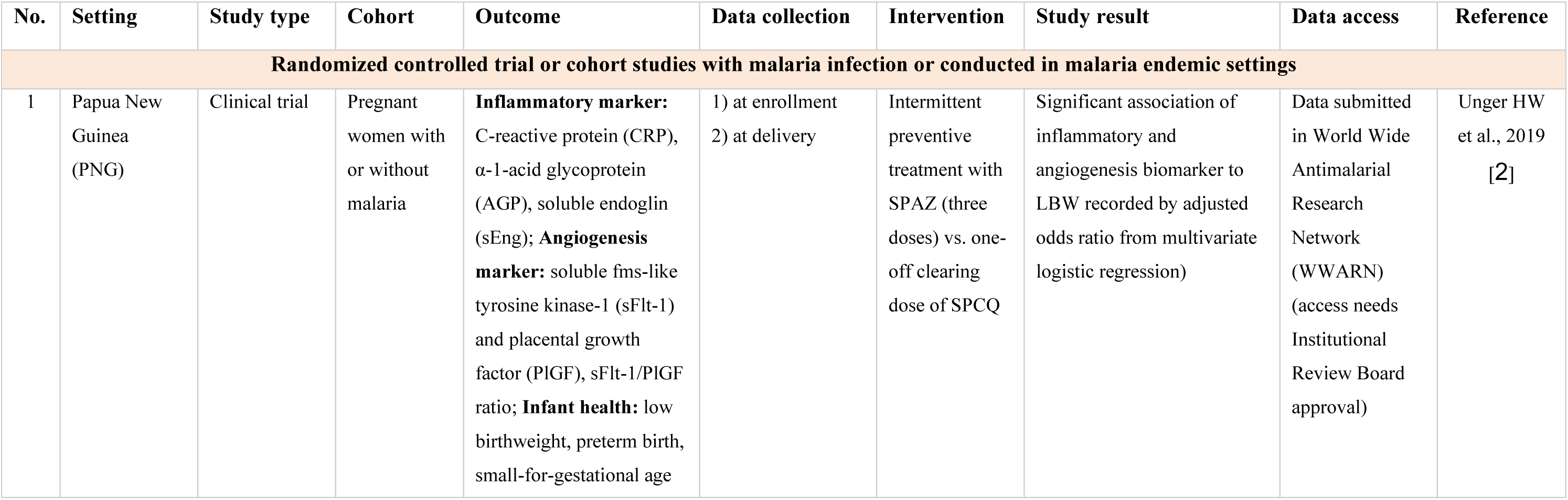
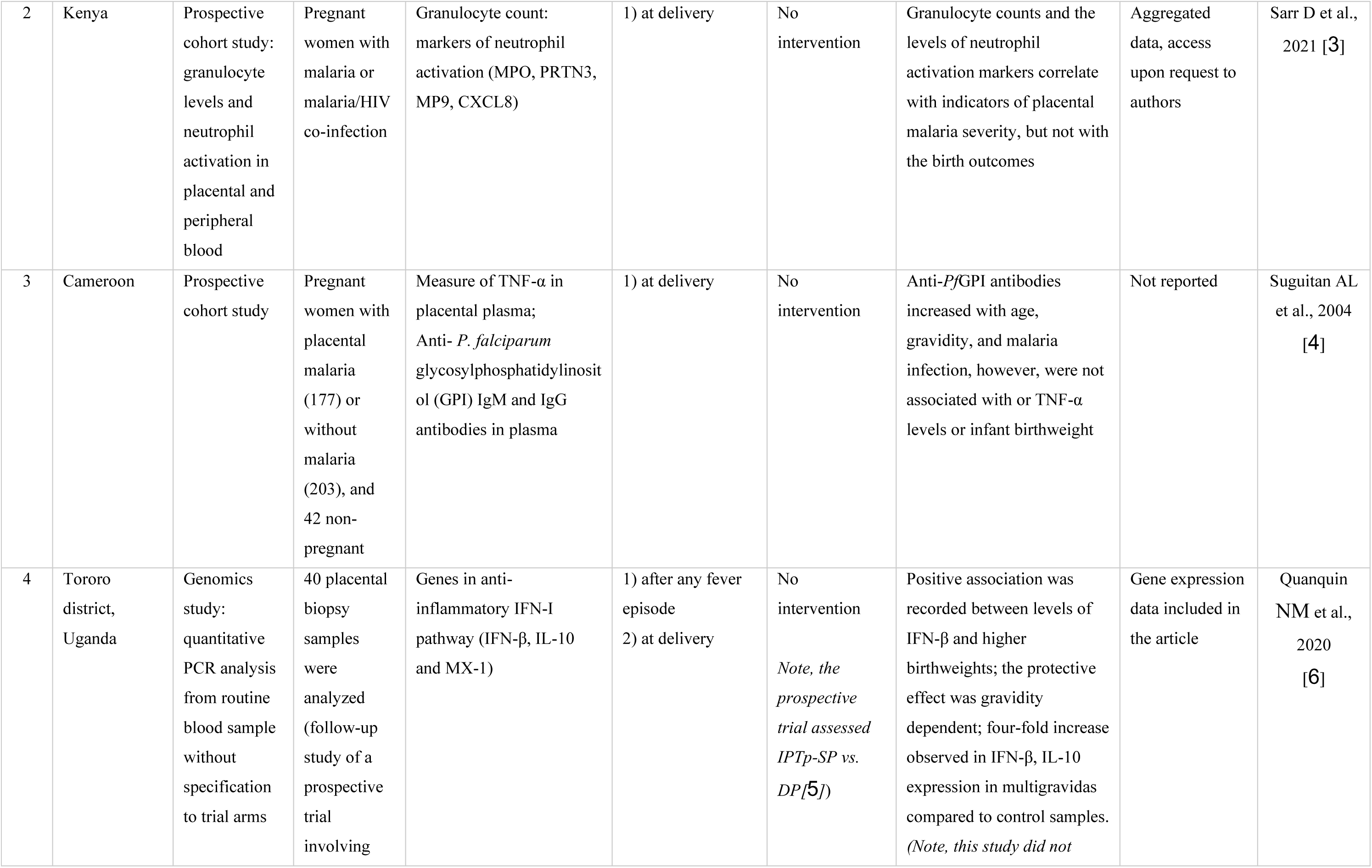
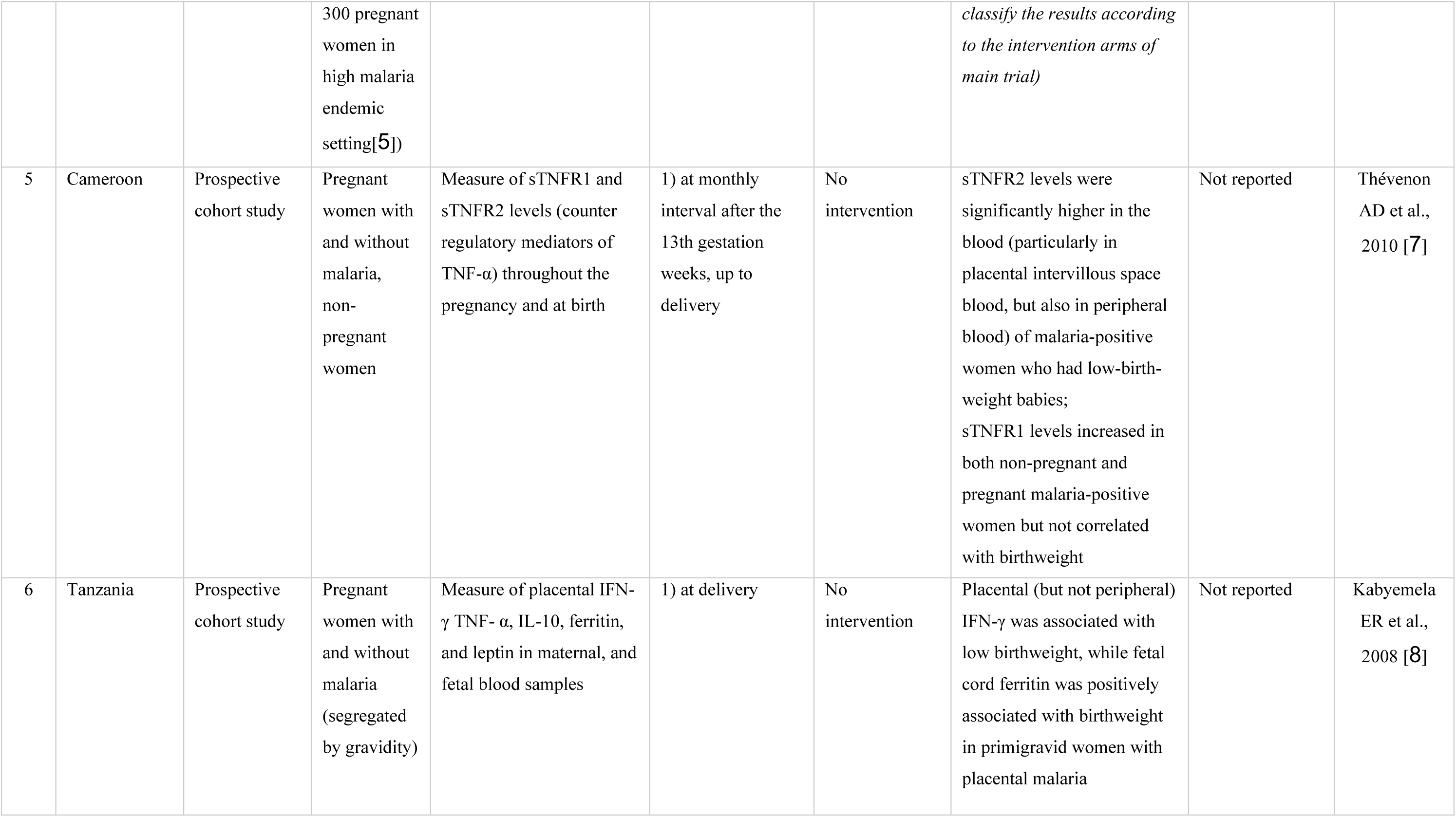
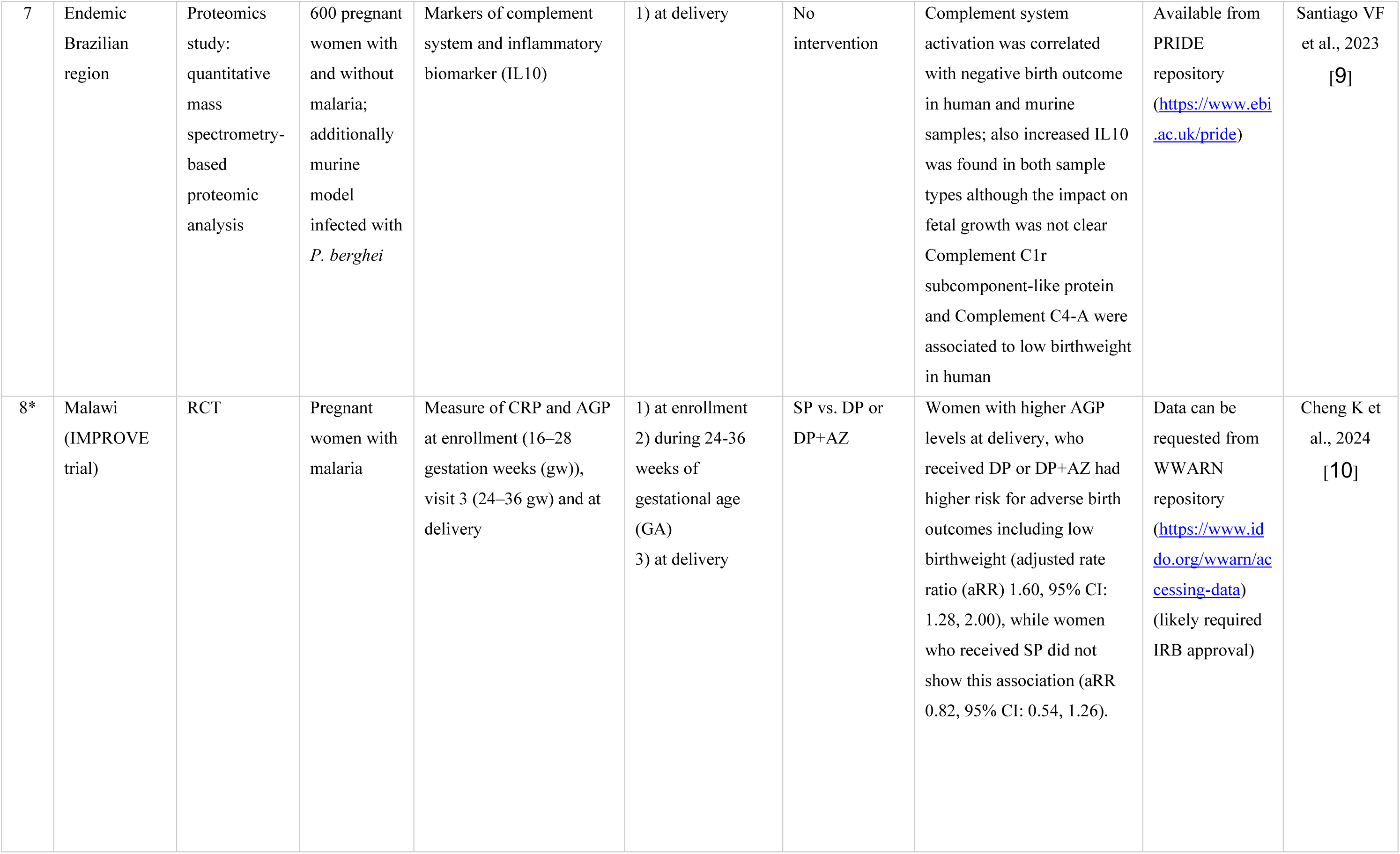
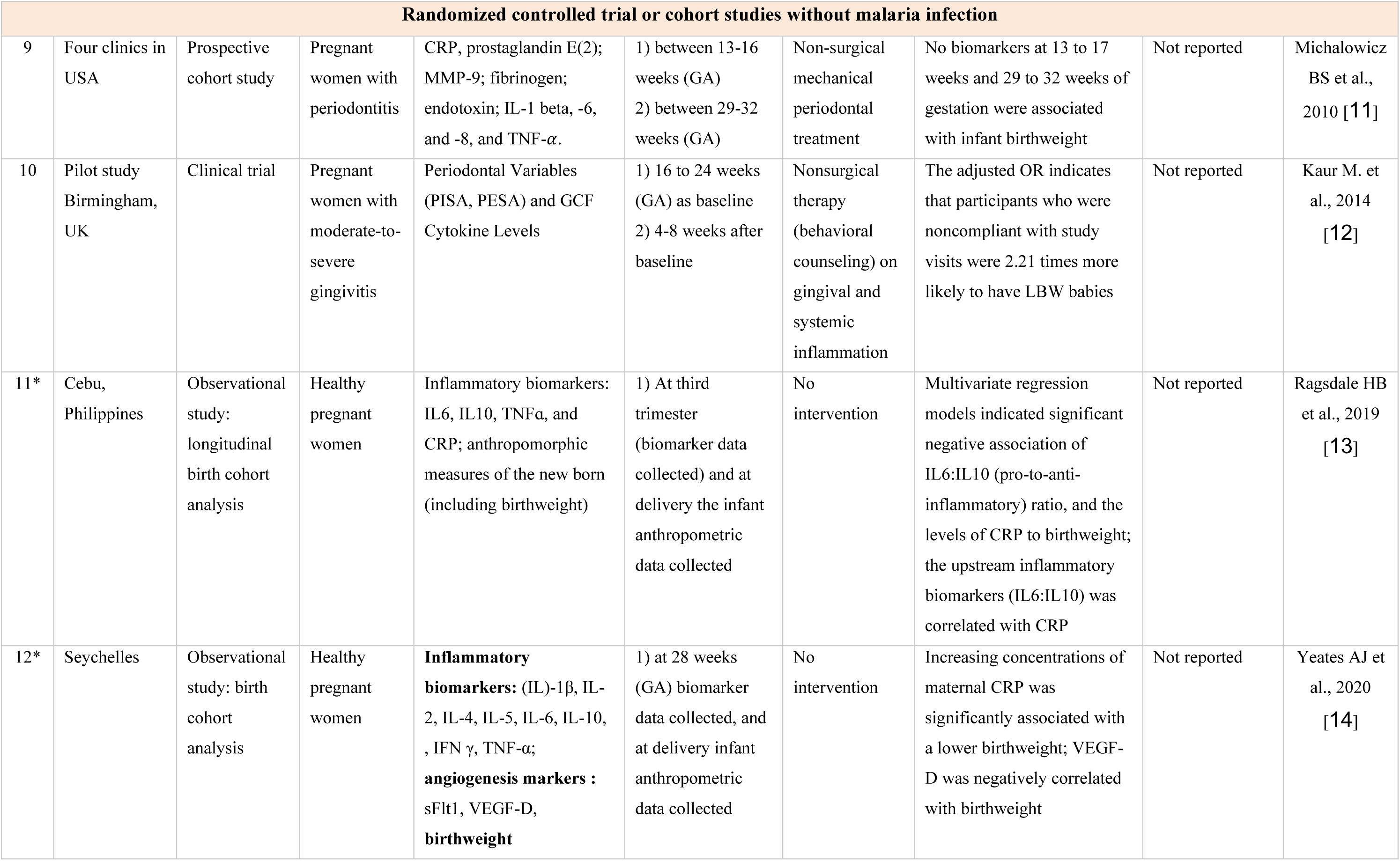
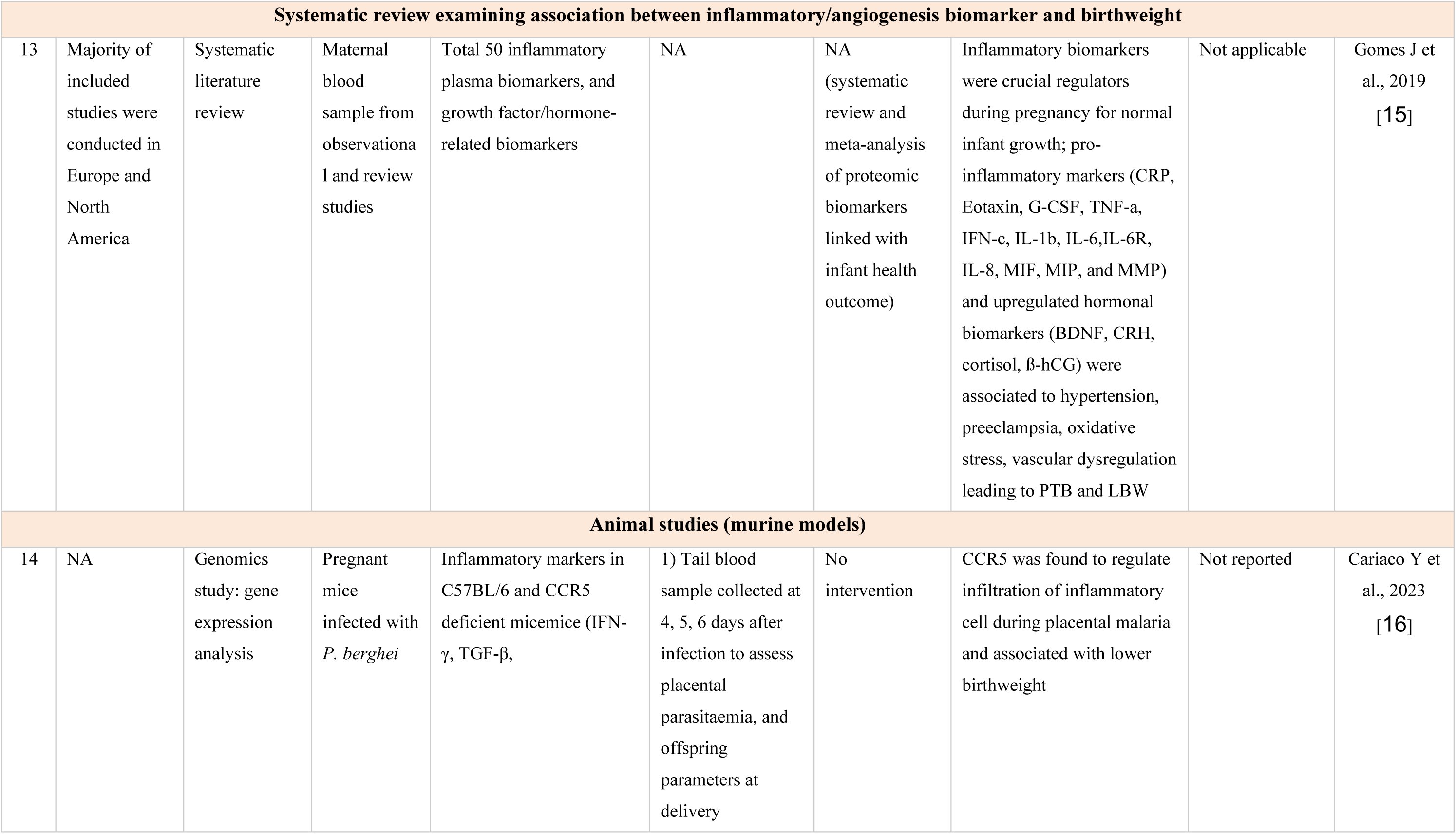
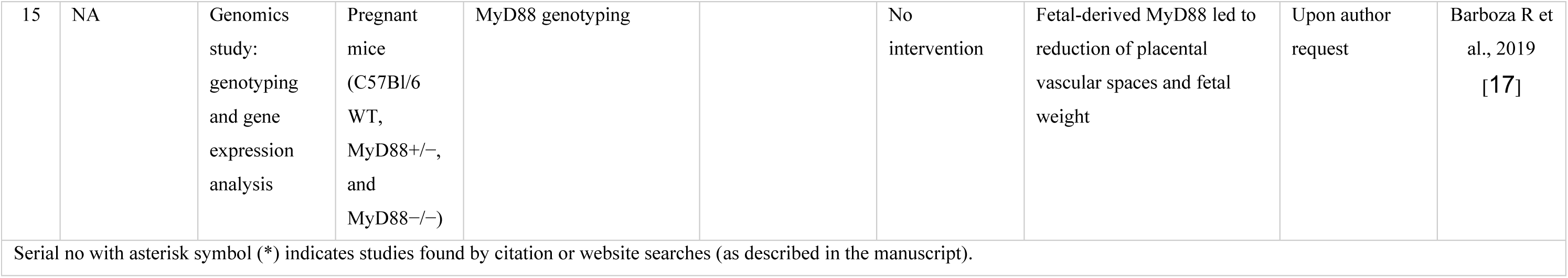
Summary of charted data from included studie.

